# Genetic insights into the association between inflammatory bowel disease and Alzheimer’s disease

**DOI:** 10.1101/2023.04.17.23286845

**Authors:** Lu Zeng, Charles C. White, David A. Bennett, Hans-Ulrich Klein, Philip L. De Jager

## Abstract

Myeloid cells, including monocytes, macrophages, microglia, dendritic cells and neutrophils are a part of innate immune system, playing a major role in orchestrating innate and adaptive immune responses. Both Alzheimer’s disease (AD) and inflammatory bowel disease (IBD) susceptibility loci are enriched for genes expressed in myeloid cells, but it is not clear whether these myeloid risk factors are shared between the two diseases. Leveraging results of genome-wide association studies, we investigated the causal effect of IBD (including ulcerative colitis (UC) and Crohn’s disease (CD)) variants on AD and its endophenotypes. Microglia and monocyte expression Quantitative Trait Locus (eQTLs) were used to examine the functional consequences of IBD and AD variants. Our results revealed distinct sets of genes and pathways of AD and IBD susceptibility loci. Specifically, AD loci are enriched for microglial eQTLs, while IBD loci are enriched for monocyte eQTLs. However, we also found that genetically determined IBD is associated with a protective effect against AD (p<0.03). Yet, a genetic propensity for the CD subtype is associated with increased amyloid accumulation (beta=7.14, p-value=0.02) and susceptibility to AD. Susceptibility to UC was associated with increased deposition of TDP-43 (beta=7.58, p-value=6.11×10^-4^). The relation of these gastrointestinal inflammatory disease to AD is therefore complex; while the different subsets of susceptibility variants preferentially affect different myeloid cell subtypes, there do appear to be certain shared pathways and the possible protective effect of IBD susceptibility on the risk of AD which may provide therapeutic insights.

## Introduction

Myeloid cells (i.e., monocytes, macrophages, microglia, dendritic cells and neutrophils), are an important component of the innate immune system and also play critical roles in tissue development, homeostasis, disease, and repair. Their regulatory architecture is gradually emerging (1, 2) and includes a number of different transcriptional programs, many of which are specialized for the tissue context in which a given myeloid cell finds itself. Dysfunction in the immunoregulatory roles of these cells may play a part in the pathogenesis of a subset to patients with inflammatory bowel disease (IBD). IBD, including its two major subtypes of ulcerative colitis (UC) and Crohn’s disease (CD), is a chronic gastrointestinal inflammatory disease whose etiology remains poorly understood and leads to recurrent attacks of acute inflammation over more chronic changes (3). Genetic studies have noted a propensity for IBD variants to influence gene expression in monocytes (4, 5), consistent with the observations that different bone marrow-derived macrophages found in the intestinal epithelium and perhaps those in the enteric nervous system contribute to these acute and chronic inflammatory changes (6). By contrast, microglia, the resident myeloid cells in the central nervous system (CNS), arise from primitive yolk-sac emigrants that infiltrate the developing neurectoderm that will differentiate into the central nervous system (CNS) (7). Activation of microglia in the human brain, concentrated around amyloid-beta (Aβ) plaques and neurofibrillary tangles (NFTs), is a prominent feature of Alzheimer’s disease (AD) pathology (8). AD is the most common aging-related neurodegenerative disease and is the most common cause of dementia (9), and the majority of AD susceptibility loci contain genes expressed by microglia and monocytes (8, 10, 11). Thus, both AD and IBD susceptibility appear to involve perturbation of myeloid cell function. Epidemiologic studies have further reported that patients with IBD had a more than two-fold increase in the risk of developing dementia (12).

Further, dementia was diagnosed around 7 years earlier in people with IBD than in those without (13). However, more robust evidence is needed to understand whether IBD and AD (instead of the broader diagnostic grouping of “dementia”) are truly linked.

Here, we leveraged existing IBD and AD genome-wide association studies (GWAS), and microglial and monocyte expression Quantitative Trait Locus (eQTL) datasets (11, 14, 15) to examine the extent to which functional consequences of IBD and AD risk variants may be shared or distinct. We investigated the genetic effect of IBD on AD and AD endophenotypes, and the genetic correlation among IBD, AD and other disease traits. Our results indicate that (1) Overall, IBD loci are different from AD loci, indicating separate genetic underpinnings for the two conditions that affect different aspects of myeloid function. (2) AD risk variants displayed a enrichment in microglia eQTLs; in contrast, risk variants associated with IBD and UC were enriched in monocyte eQTLs, highlighting the context-specific effects of these variants. (3) Genetically determined IBD was consistently linked to a reduced risk of AD, suggesting a protective genetic influence. (4) Conversely, a propensity for the IBD subtype of CD was associated with a higher risk of AD, possible due to its positive effect on the accumulation of amyloid within cerebral blood vessles, and genetic susceptibility for UC was found associated with increased TDP-43 burden. Thus, we find that these two clinically distinct entities – IBD and AD – have important contributions from myeloid cells in the early functional consequences of susceptibility variants that are largely distinct even if they affect the same cell types; nonetheless, there may be a modest genetic interaction between these two groups of susceptibility variants.

## Materials and methods

### IBD and AD variants collection

A recent study reported a combination of 241 known SNPs with the smallest p-value in fifteen studies associated with IBD/UC/CD (16). We extracted the GWAS summary statistics (e.g., effect allele, effect size and p-value) from this study, p-values smaller than 5×10^-8^ were retained for the following analyses, resulting in 81 IBD risk variants, 18 CD risk variants and 15 UC risk variants (**Supplementary Table S1**).

For AD variants, previous study identified 38 independent AD risk loci from 90,338 cases (46,613 proxy) and 1,036,225 controls (318,246 proxy) (17), which were all reached genome-wide significance, with p ≤ 5×10^-8^ were kept (**Supplementary Table S2**).

### AD endophenotypes

Ten AD endophenotypes were used in omnibus test and Phenome-wide association studies (PheWAS) analysis, which were AD pathologies measurement from 2,409 ROSMAP participants (18) (details can be seen in **Supplementary Table S3**). For instance, cogng_demog_slope, is the random slope of global cognition that estimated person-specific rate of change in the global cognition variable over time. It comes from a linear mixed effects model with global cognition as the longitudinal outcome. The model controls for age at baseline, sex, and years of education. amyloid_sqrt is the amyloid beta protein identified by molecularly-specific immunohistochemistry and quantified by image analysis. Value is percent area of cortex occupied by amyloid beta. Mean of amyloid beta score in eight regions (https://www.radc.rush.edu/docs/var/variables.htm). For these 2,409 ROSMAP participants, their age at death ranged from 66yrs to 108yrs, with an average of 89.5yrs (SD=±6.53), among them 718 were male, and 1,691 were female.

### eQTL data

Monocyte cis-eQTL was downloaded from EMBL-EBI eQTL catalogue (https://www.ebi.ac.uk/eqtl/) (14), which aims to provide uniformly processed gene expression from all available public studies on human. The monocyte RNA-seq data was originally from BLUEPRINT study (https://www.blueprint-epigenome.eu/) (15), based on 197 European individuals, generated from whole blood. More details can be found at https://www.ebi.ac.uk/eqtl/Methods/.

Microglia single-nucleus RNA sequencing data was generated from the dorsolateral prefrontal cortex (DLPFC) of 424 ROSMAP (the Religious Order Study and the Rush Memory Aging Project). Both studies are harmonized and include older individuals tested annually all of who agreed to brain donation. They were approved by an Institutional Review Board of Rush University Medical Center. All participants signed informed and repository consents, and an Anatomical Gift Act. Matrix eQTL (19) version 2.3 method was then used to identify cis-eQTLs with 1 MB of the transcription start site of each measure gene. More details can be found in our recent work (11). In this study, we only kept the eQTL-eGene pairs with Bonferroni-Hochberg (BH) p-value<0.05, resulting in 925,472 monocyte eQTL-eGene pairs, and 100,578 microglia eQTL-eGene pairs.

### Colocalization analysis

The COLOC package (version 5.1.0) was applied to test the approximate Bayes factor (ABF) colocalization hypothesis, which assumes a single causal variant. Under ABF analysis, the association of a trait with a SNP is assessed by calculating the posterior probability (value from 0 to 1), with the value of 1 indicating the causal SNP. In addition, the ABF analysis has 5 hypotheses, where, PP.H0.abf indicates there is neither an eQTL nor a GWAS signal at the loci; PP.H1.abf indicates the locus is only associated with the GWAS; PP.H2.abf indicates the locus is only associated with the eQTL; PP.H3.abf indicates that both the GWAS and eQTL are associated but to a different genetic variant; PP.H4.abf indicates that the eQTL and the GWAS are associated to the same genetic variant. With the posterior probability of each SNP and aiming to find the casual variants between the GWAS and eQTL, we focused on extracting the PP.H4 value for each SNP in our study.

For GWAS studies, we used the reported lead SNPs for each trait. For example, 38 AD risk loci, 81 IBD risk variants, 19 CD risk loci and 15 UC risk variants as we mentioned above. For each locus, we searched for the eSNPs that are within 500 kb of the lead SNP, and listed eGenes that were paired with the eSNP in microglia and monocyte eQTL datasets (15, 20). We then obtained the eGenes cis-eQTL output around the lead eSNP within 1 Mbp window size. In addition, we extracted GWAS summary statistics around the reported IBD/CD/UC lead SNP. At last, we conducted COLOC for respective pair of eGene-eQTL and eSNP-GWAS.

### Omnibus test

PLINK/1.9 –score (21) was used to calculate the polygenic risk score (PRS) of 81 IBD/19 CD/15 UC risk on 2,409 ROSMAP donors. We then applied a generalized linear regression model using the R “glm” function to conduct the association test between the impact of IBD/CD/UC risk on ten AD endophenotypes (details can be seen in **Supplementary Table S3**). Adjusting for the covariates of age at death, sex and education years.

### PheWAS analysis

Phenome-wide association studies (PheWAS) was further used to test the associations between 81 IBD/19 CD/ 15 UC risk variants (exposure) and ten AD endophenotypes (outcome), using the R PheWAS package (22), adjusting for age at death, sex and education years. We used a linear regression model giving the numeric phenotype outcomes. We established significance using an FDR corrected *P-*value, accounting for ten AD endophenotypes.

### Mendelian randomization

We performed Generalized Summary-data-based Mendelian Randomization (GSMR) (23) analyses between IBD (exposure), including CD (exposure) and UC (exposure) on AD (outcome) of European descent using GWAS summary statistics, implemented in GCTA software (v1.93.1f beta). The instrumental variables were selected by a clumping procedure internal to the GSMR software with parameters: --gwas-thresh 5 × 10^−8^ --clump-r2 0.05. The HEIDI-outlier test was applied to test for horizontal pleiotropy (*P*_HEIDI_ < 0.01). For comparison, we performed two-sample MR analysis (2SMR) with the inverse-variance-weighted regression model and Egger regression model, implemented via the TwoSampleMR R package (24).

### LD score regression (LDSC)

LDSC (25) bivariate genetic correlations attribute to genome-wide SNPs (r_g_) were estimated with eleven human disease traits from published GWASs, including three IBD associated traits (IBD (*N_cases_* =25,042, *N_controls_*=34,915), CD (*N_cases_* =12,194; *N_controls_*=28,072) and UC ((*N_cases_*=12,366; *N_controls_*=33,609)) (16), three neurodegenerative disorders: AD (*N_cases_*=90,338; *N_controls_*=1,036,225) (17), ALS (*N_cases_*=27,205; *N_controls_*=110,881) (26) and Parkinson’s disease (PD) (*N_cases_*=37,688; *N_cases_proxy_*= 18,618; *N_controls_*=1,400,000) (27), another inflammatory diseases, multiple sclerosis (MS) (*N_cases_*=47,429; *N_controls_*=68,374) (28) and a metabolic disease type 2 diabetes (T2D) (*N*_cases_=77,418; *N*_controls_=356,122) (29) and three psychiatric disorders: bipolar disorder (BD) (*N*_cases_=41,917; *N*_controls_=371,549) (30), major depressive disorder (MDD) (*N*_cases_=170,756; *N*_controls_=329,443) (31) and schizophrenia (SCZ) (*N*_cases_=76,755; *N*_controls_=243,649) (32). Adjusting for the number of traits tested, the FDR *P*-value threshold (FDR<=0.05) was used to define significant genetic correlations.

## Results

### Cell state specificity of IBD and AD

To understand the functional consequences and potential interactions of IBD and AD susceptibility loci, we compiled a list of 81 IBD risk variants, 19 CD risk variants and 15 UC risk variants (meeting a threshold of genome-wide significance, p≤5×10^-8^) reported in a recent GWAS(16). In addition, we included 38 AD risk variants meeting the same threshold from a recent AD GWAS (17) (**Supplementary Table S1 & S2**) (**Fig. 1A**). We first investigated whether there was any overlap between the two lists of susceptibility loci, and our results showed no direct overlap between the top SNPs in the IBD and AD.

**Fig. 1.**
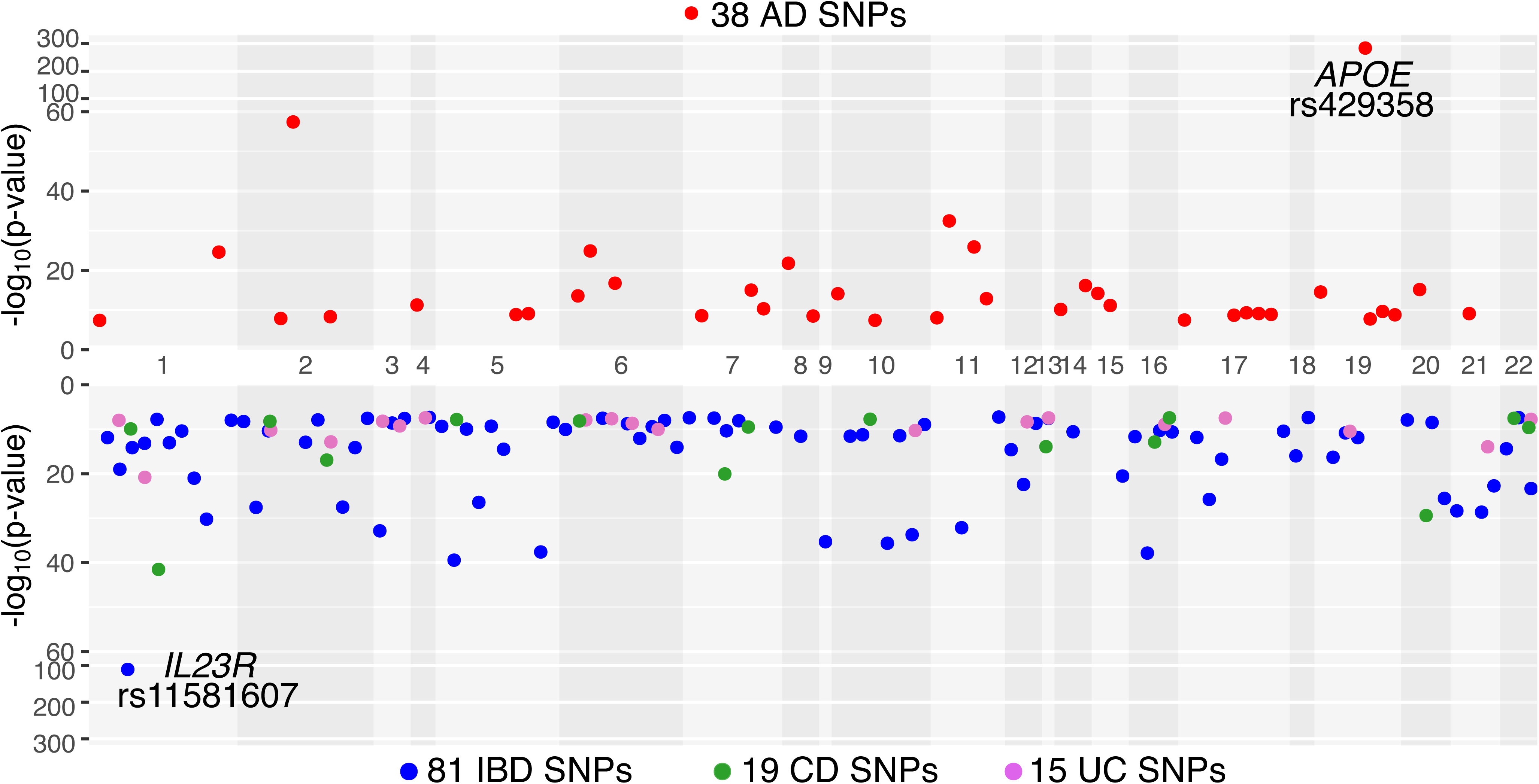
Overview of AD and IBD risk variants used in this study. **A)** Manhattan plot of 38 genome-wide significant (p<=5 × 10^-8^) AD risk variants and 81/19/15 genome-wide significant (p<=5 × 10^-8^) IBD/CD/UC risk variants extracted from previous studies.

IBD and AD susceptibility loci have been reported to be enriched for genes expressed in peripheral monocytes (10, 33, 34), which are mostly recent bone marrow immigrants that are relatively undifferentiated. Myeloid cells adopt their final, functional states in tissue, in response to the local environment, and thus we assessed whether the microglial state of differentiation would yield differential enrichment for AD loci *vs.* the monocyte state. To do this, we downloaded monocyte eQTL results (15) from the EMBL-EBI eQTL catalogue (14) (available at https://www.ebi.ac.uk/eqtl/) and used microglia eQTLs sourced from our recent study (20). In this study, we only kept the eSNP-eGene pairs with BH adjusted p-value<0.05, resulting in 925,472 monocyte eSNP-eGene pairs, and 100,578 microglia eSNP-eGene pairs. Integrating these results with the disease susceptibility results show that >21.1% of AD (8/38) lead SNPs enriched among microglia eQTLs (hypergeometric p-value = 1.17×10^-8^). On the other hand, <2.5% of IBD (2/81), CD (0/19), and UC (0/15) are microglia eSNPs (**Supplementary Table S4**). Different from microglia, >19.7% of AD, IBD, CD and UC were monocyte eSNP (AD: hypergeometric p-value = 2.18×10^-5^, IBD: hypergeometric p-value = 5.20×10^-4^, CD: hypergeometric p-value = 0.06, UC hypergeometric p-value = 0.03) (**Supplementary Table S4**). Some of the difference in overlap with AD loci between the monocyte and microglial eQTLs is likely to be due to the parent studies given the smaller sample size and nuclear RNA-derived nature of the microglial data; both of these factors will tend to diminish the number of eQTL discovered.

Integrating microlgia and monocyte eQTLs with the disease GWAS results, we uncovered evidence of colocalization (PP.H4>0.8) for nine eGenes among nine AD loci within microglia (**Fig. 2A & Supplementary Table S5**). We confirm some of the well-validated results, such as *BIN1, USP6NL* and *PICALM*. These microglial eQTLs colocalized with AD susceptibility were absent in monocytes (35). On the contrary, only one eQTL target gene was found to be colocalized with IBD risk variants (**Fig. 2B**) in microglia. ETS Proto-Oncogene 2 (*ETS2*), which plays a pivotal role as a transcription factor driving inflammation in acutes and chronic inflammatory disease, upregulation of genes with ETS2-binding sites has been previously reported in UC patients (36).

**Fig. 2.**
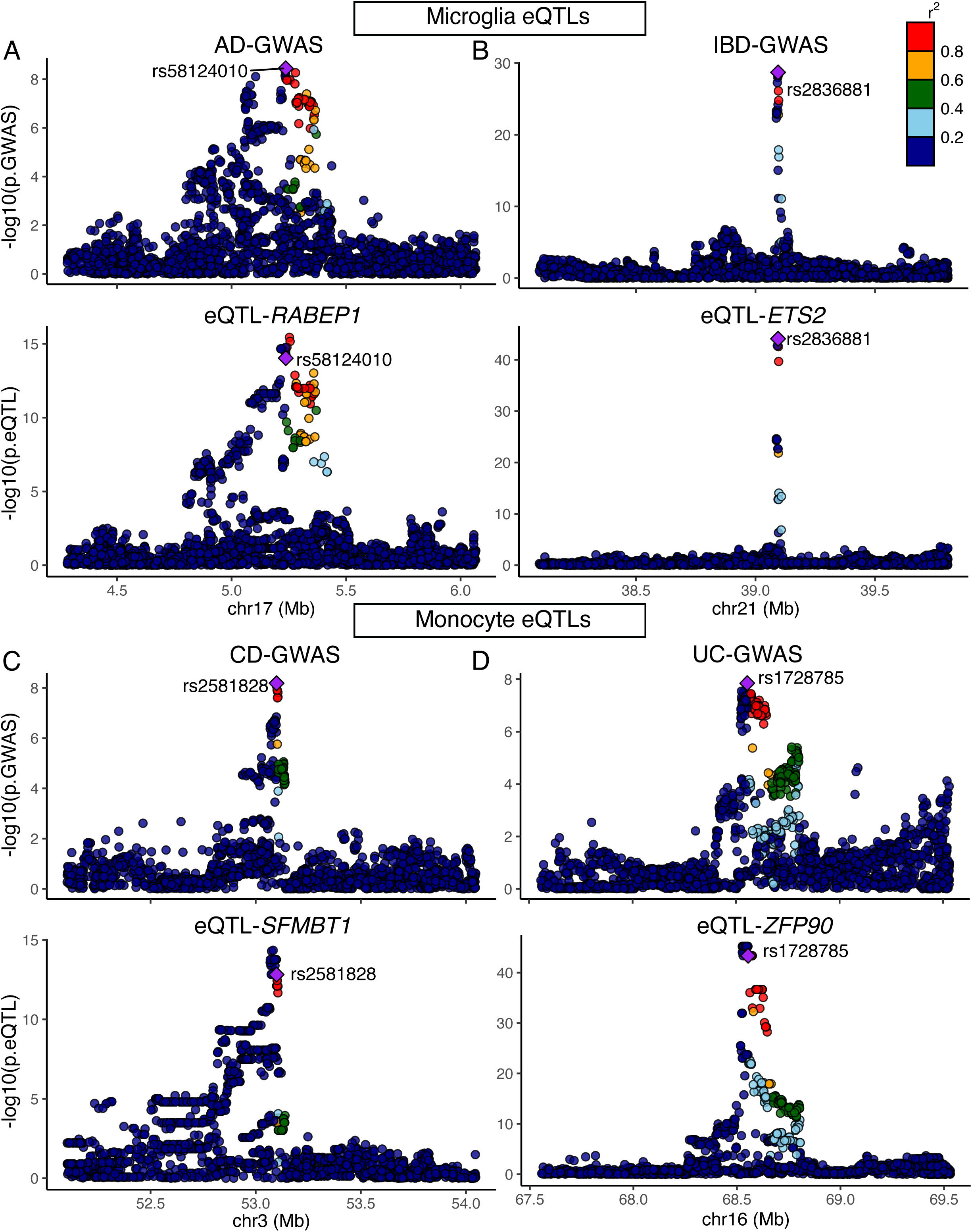
Colocalization between IBD/UC/CD/AD risk variants and microglia/monocyte eQTLs. A) Example of eQTL-AD-GWAS colocalization near the *RABEP1* locus in microglia. B) Example of eQTL-IBD-GWAS colocalization near the *ETS2* locus in microglia. C) Example of eQTL-CD-GWAS colocalization near the *SFMBT1* locus in monocyte. D) Example of eQTL-UC-GWAS colocalization near the *ZFP90* locus in monocyte.

With respect to monocyte eQTLs, only two eGenes showed colocalization evidence in AD, they are *APH1B* and *MADD*. Dysregulated expression levels of *APH1B* in peripheral blood were reported associated with brain atrophy and amyloid-β deposition in Alzheimer’s disease (37). However, we identified seven monocyte eQTL effects colocalized with five IBD risk loci, three colocalized with two CD risk loci, and five colocalized with five UC risk loci (**Fig. 2C & D, supplementary Table S5**), For example, Scm Like With Four Mbt Domains 1 (*SFMBT1*) is a polycomb protein with transcriptional repressor activity (38), and was identified as target genes for IBD-associated variatns (39). ZFP90 Zinc Finger Protein (*ZFP90*), which has been reported to drive the initiation of colitis-associated colorectal cancer (40). The dysregulation of *ZFP90* was associated with some autoimmune diseases, including inflammatory bowel disease, vitiligo, and multiple sclerosis (41).

### Genetic architecture of IBD and AD

We then examined the impact of 81 genome-wide significant IBD risk loci on AD- and aging-related pathological features by aggregating the effect of these variants into a wighted genetic risk score (GRS) and then conducting an omnibus test, adjusting for age at death, sex and education, which evaluates the distribution of the associations with ten pathologic endophenotypes (details can be seen in **Fig. 3** legend and **Supplementary Table S3**). In this omnibus test, the distribution of results for the individual pathologic traits is not significant, and none of the individual traits reaches a threshold of nominal sifnificance (**Fig. 3A**). Expanding our analysis, we explored a CD GRS and a UC GRS; the omnibus analyses for these GRS was also non-significant. However, we noted a nominally significant (p-value<0.05) effect on cerebral amyloid angiopathy (CAA) accumulation (p-value=0.02) (**Fig. 3B**), and the UC GRS demonstrated a significant association with the extent of TDP-43 deposition (p-value=6.11×10^-4.^, FDR=6.11×10^-3^) (**Fig. 3C**). The latter result is the more intriguing one given its significance, and the fact that the biology of TDP-43 deposition remains less well understood.

**Fig. 3.**
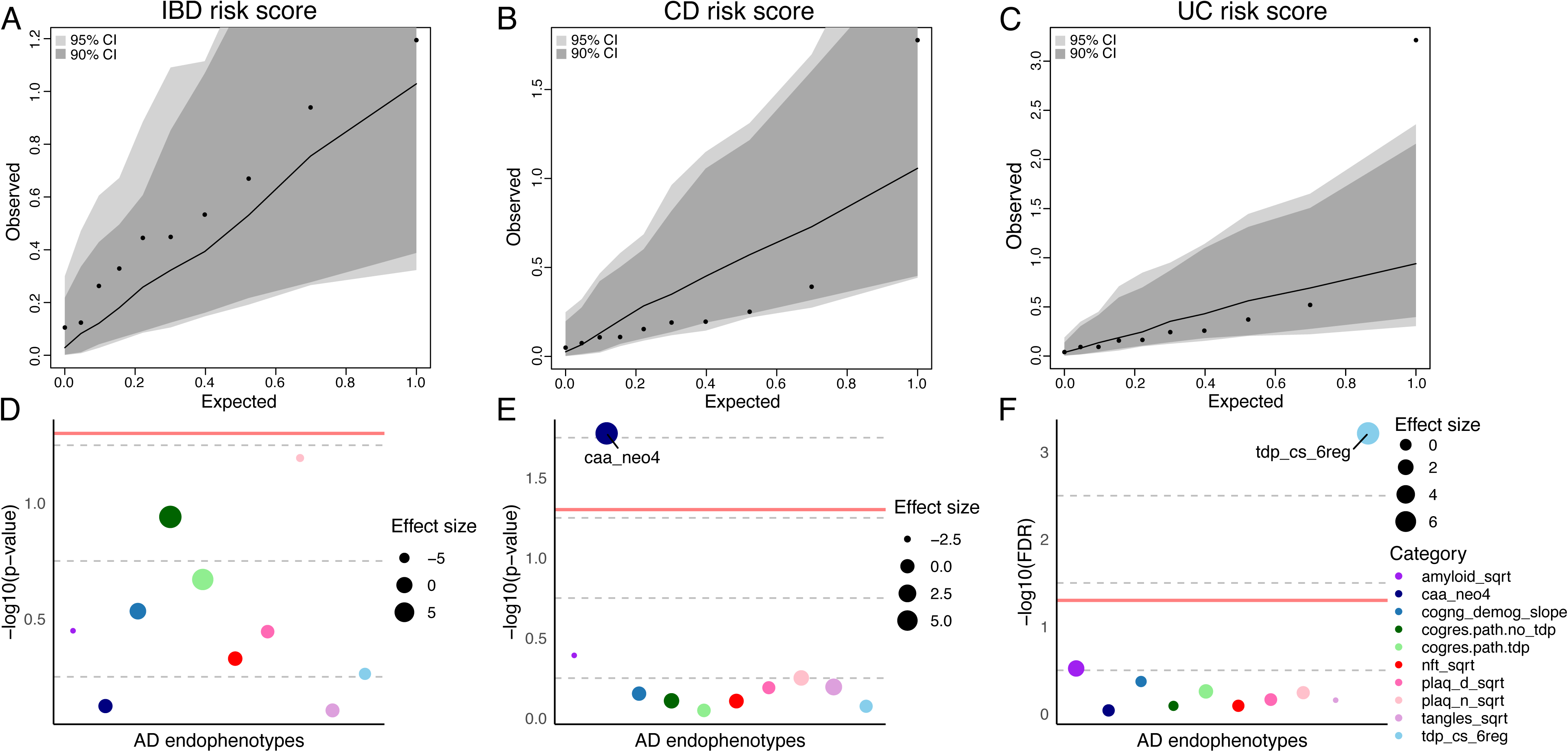
Genetic analysis between IBD and AD endophenotypes. A) The impact of IBD risk on AD endophenotypes (e.g. amyloid, plaque and tangles). Each dot represents a ROSMAP AD endophenotype and its expected and observed associations with AD endophenotype. The expected P values assume a null distribution with no linear associations between IBD risk score and AD pathologies after adjusting age at death, sex and education. The gray and dark areas indicate the extreme ranges of the QQ plot as generated by chance at a threshold of *P*=0.05 and *P*=0.10, respectively. The 95% and 90% confidence intervals were empirically derived by randomly assigning participants with the IBD risk score and repeating the analysis 1,000 times. Based on the distribution of the observed *P* values for all the ten AD phenotypes. B) Phenome-wide association plots of IBD risk with the ROSMAP AD endophenotypes. Each dot represents a ROSMAP AD endophenotypes, the red line is the p-value threshold (p≤0.05). The effect size denotes the beta value.

To complement these analyses, we deployed a Phenome-wide association study (PheWAS) approach to explore the associations between the IBD/CD/UC variants (exposure) and ten AD endophenotypes (outcome) using the R PheWAS package (22), adjusting for age at death, sex and education. Employing a linear regression model to accommodate numeric phenotype outcomes, our results revealed that IBD had no significant effect on AD endophenotypes (**Fig. 3D**). While CD displayed a nominally significant impact on cerebral amyloid angiopathy (beta=7.14, p-value=0.02) (**Fig. 3E**), and UC demonstrated a notable effect on TDP-43 severity (beta=7.58, p-value=6.11×10^-4^, FDR=6.11×10^-3^) (**Fig. 3F**). Based on the analyses above, our findings suggest that CD risk loci may have a small contribution to CAA risk, whereas UC risk loci may have an effect on TDP-43, consistent with the earlier analyses.

Subsequenctly, we utilized generalized summary statistics-based Mendelian randomization (GSMR) (23) and two-sample Mendelian randomization (2SMR) (24) to investigate the causal impact of IBD (exposure, *N_cases_* =25,042; *Ncontrols=*34,915), CD (exposure, *N_cases_* =12,194; *Ncontrols=*28,072) and UC (exposure, *N_cases_*=12,366; *N_controls_*=33,609) on AD (outcome, *N_cases_*=90,338; *Ncontrols=*1,036,225) among individuals of European descent. Among these risk variants, 78 out of 81 were found shared between IBD and AD summary statistics, 19 out of 19 were common between CD and AD summary statistics, and 15 out of 15 variants overlapped between UC and AD summary statistics.

The results of mendian randomization indicated that genetically determined IBD was significantly associated with a decreased susceptibility of AD (GSMR: b_xy_=-4.76×10^-3^, p-value=9.48×10^-3^, Inverse variance-weighted (IVW): b_xy_=-5.69×10^-3^, p-value=0.03) (**Fig. 4A & D**). However, our subanalyses indicated an increased causative relationship betweeen CD and AD (GSMR: b_xy_=9.83×10^-3^, p-value=0.02, IVW: b_xy_=0.01, p-value=0.02, weighted mean: b_xy_=0.01, p-value=0.01) (**Fig. 4B & E**), while UC exhibited no causal associations with AD (**Fig. 4C & F**) (**Supplementary Table S6**).

**Fig. 4.**
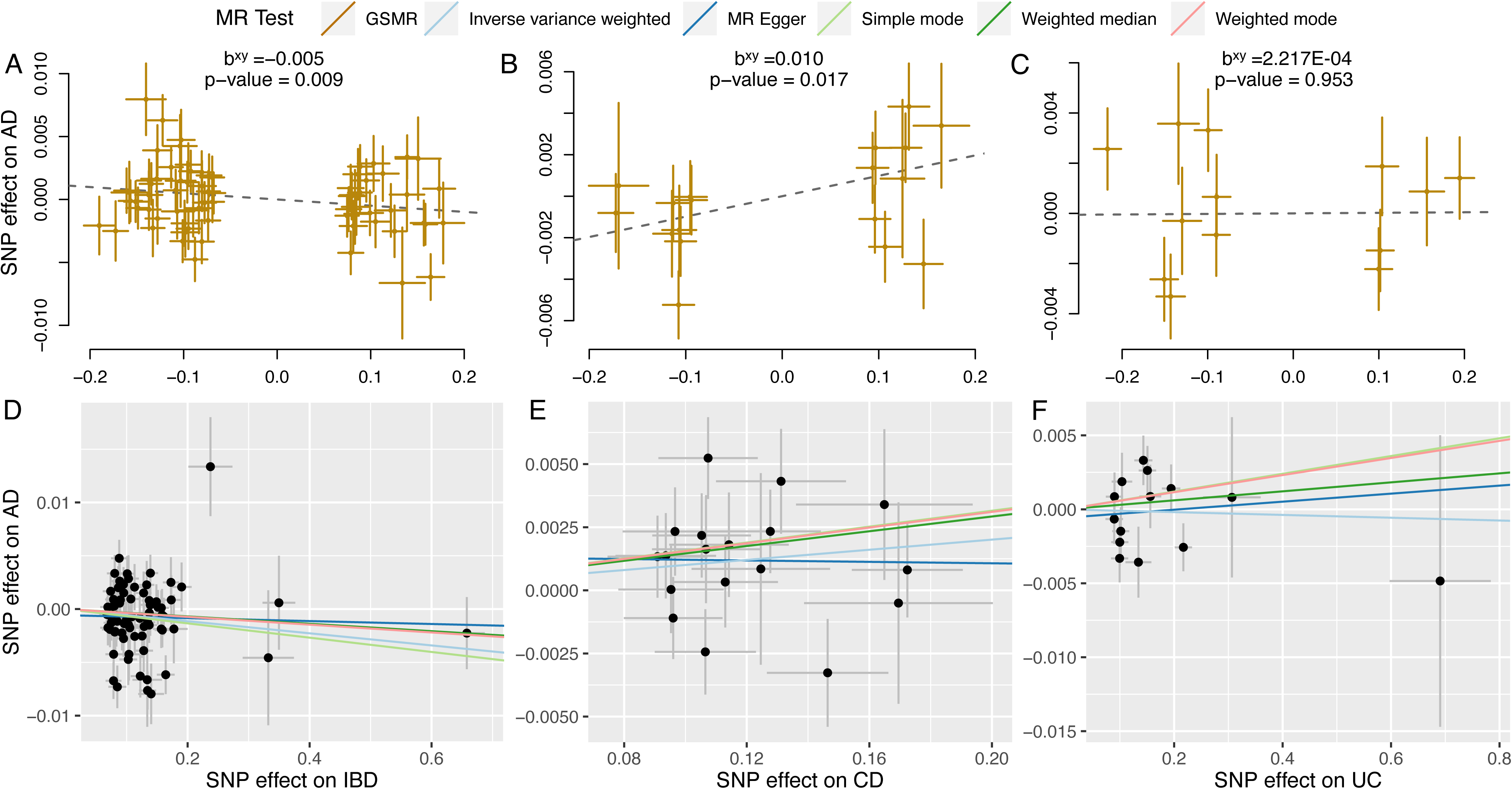
Scatter plots of MR analyses showing the causal effects of IBD on AD. Six different methods, including: Generalised Summary-data-based Mendelian Randomisation (GSMR), default inverse variance-weighted (IVW), weighted mode, weighted median, Mendelian randomization-Egger (MR-Egger) regression and simple median were used to evaluate the causal effects of IBD (UC and CD) on AD. The slope in each line represents the causal estimate of an exposure on corresponding outcome per method. (A, D) SNP’s effect on IBD and AD; (B, E) SNP’s effect on CD and AD; (C, F) SNP’s effect on UC and AD.

### Genetic correlation between IBD and other traits

Finally, we employed LD score regression (LDSC) (25) to estimate genetic correlations between IBD and AD, along with other disease traits. To do this, we downloaded GWAS summary statistic from nine previously published studies, which were all found to be enriched for loci encoding myeloid genes. Including three IBD traits (IBD, CD and UC) (16), three neurodegenerative disorders (AD (17), ALS (26) and Parkinson disease (PD) (27), another inflammatory disease (Multiple sclerosis, MS (28)), a metabolic disease, type 2 diabetes (T2D) (29) and three psychiatric disorders: bipolar disorder (BD) (30), major depressive disorder (MDD) (31) and schizophrenia (SCZ) (32). **Fig. 5** inllustrates genetic correlations for all 55 pairwise combinations of these eleven traits. To define significant genetic correlations, we adjusted for the number of traits tested, using an FDR-corrected P-value threshold (FDR≤0.05).

**Fig. 5.**
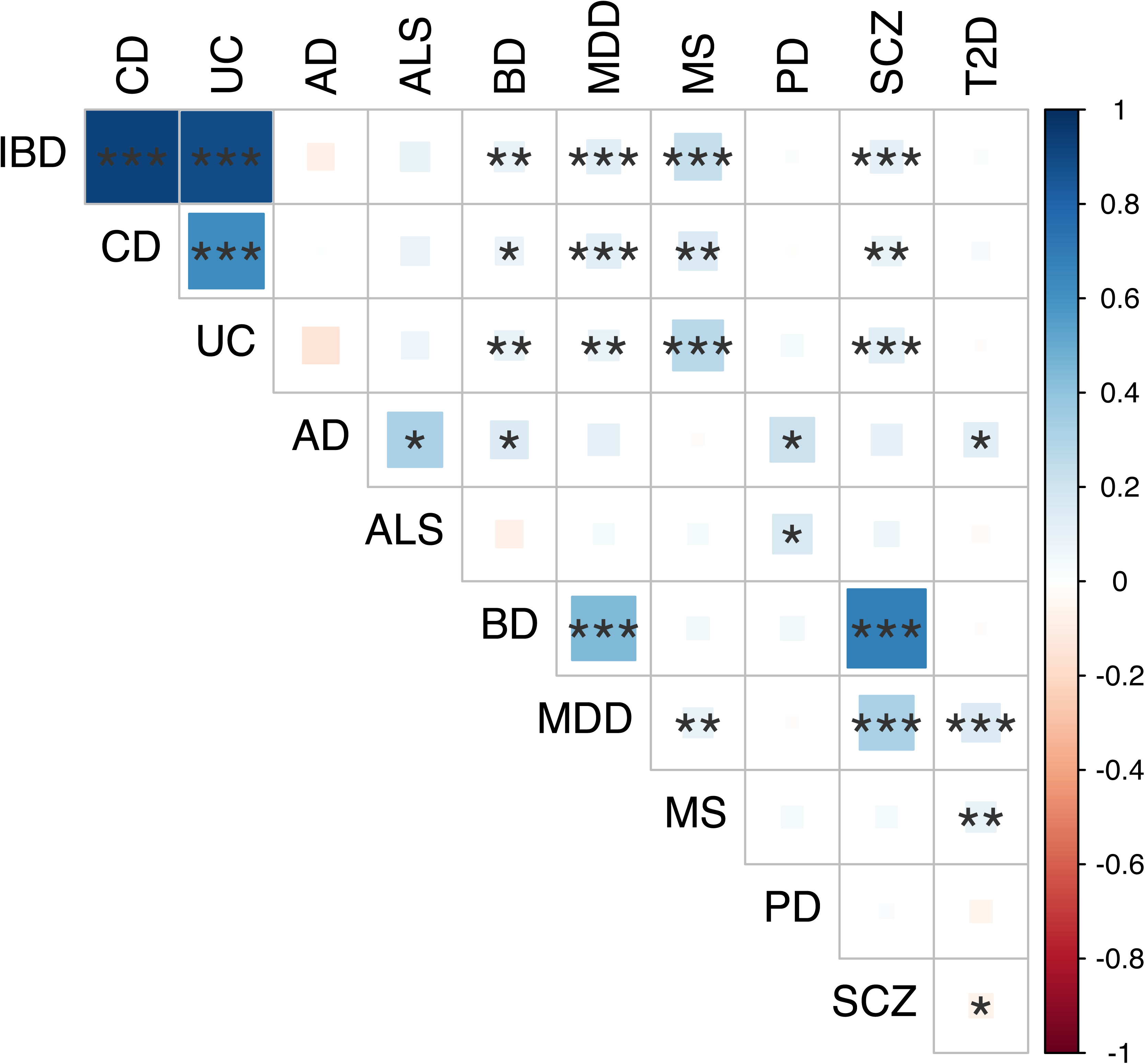
Genetic correlation between neurological and inflammatory diseases. Genetic correlation estimates across 11 human diseases. The areas of the squares represent the absolute value of corresponding genetic correlations. After FDR correction at 5% significance level, genetic correlations estimate that are significantly different from zero are marked with an asterisk (* 0.01<FDR<=0.05; ** 0.001 <FDR<=0.01; *** FDR<=0.001). The blue color denotes a positive genetic correlation, the red color denotes a negative genetic correlation.

Based on LDSC analysis, IBD-related traits showed no significant genetic correlation with AD, ALS, PD and T2D. However, they displayed positive correlations with MS (FDR<=6.69×10^-3^), as expected given prior reports (42)), as well as with all psychiatric disorders (FDR<=0.01). In these analyses, we found AD exhibited modest positive genetic correlations with ALS (FDR=0.01), BD (FDR=0.04), PD (FDR=0.03), and T2D (FDR=0.04) (**see Supplementary Table S7**).

## Discussion

Our study systematically assessed the potential shared genetic architecture between IBD and AD, given that both sets of susceptibility loci display an enrichment for susceptibility genes expressed in myeloid cells. Such shared architecture could significantly accelerate AD studies given the large number of IBD loci. However, we found that, overall, AD risk variants are different from IBD risk variants, and AD variants involve functional consequences that are primarily found in the the CNS-specific myeloid context of microglia (43). Microglia are the resident macrophages of the CNS, they play key roles in brain development and physiology during life and aging. By contrast, patients with IBD reportedly have peripheral “monocytosis”, an excess of monocytes that has been proposed as a biomarker of severity in IBD (4); this cell type harbors several of the functional cosnequences of IBD variants. Microglia and monocytes are ontogenetically distinct: while they are both derived from the mesoderm, microglia derived from yolk-sac progenitors during embryogenesis (44), whereas monocytes continuously differentiate throughout postnatal life from bone marrow hematopoietic stem cells. Distinct developmental origin and renewal mechanisms imply that monocyte-derived macrophages and microglia-derived macrophages that infiltrate tissue, including the brain, may exert different functions in pathological processes.

While our examination of individual susceptibility loci revealed no sharing between AD and IBD, our findings using more global approaches highlighted modest but intriguing evidence for a genetically driven risk effect of IBD susceptibility on AD, which is consistent with earlier reports (12, 13). In contrast, another previous study found no association between the risks of neurodegenerative diseases and genetically predicted UC or CD (45), which may be due to the smaller sample size of the IBD/CD/UC study used in this research. Overall, the effect of greater CD or UC polygentic risk on cerebral amyloid angiopathy and TDP-43 severity, respectively, may provide potential mechanistic hypotheses for the development of these neuropathologic features.

The genetic correlation of IBD with MS was consistent with previous reports, suggesting that our rigorously applied methodology produce robust, expected results in the correct direction of effect (46). Our results also showed significant positive genetic associations between IBD and psychiatric disorders (BD, SCZ and MDD). Psychiatric comorbidity, including depression, anxiety, bipolar disorder, and schizophrenia, is known to be more prevalent in IBD patients (47, 48).

Our study was motivated to evaluate the possibility of shared functional consequences of AD variants in myeloid cells, which would have enabled us to accelerate the functional dissection of innate immune perturbations that contributes to these two diagnostically distinct diseases. We conclude that, while the two sets of variants target myeloid cell types, they cause largely distinct perturbations expression, and there is only modest evidence that a genetic propensity for IBD influences AD risk. The most intriguing result may be the connection of TDP-43 proteinopathy with the variants included in the UC GRS; this provides insights into this pathophysiologic feature that has pleiotropic manifestations, including connections to the ALS - Frontotemporal dementia - cognitive aging spectrum. TDP-43 was identified as the major pathological protein in sporadic ALS and in the most common pathological subtype of FTD (49).In addition, the standardized incidence ratios for daughters for UC was found 2.02 when mothes were diagnosed with ALS (50). Notely, there is a strong genetic enrichment (160-fold) observed between FTD and UC (51), highlighting the complex interplay between these conditions and underscoring the need for more comprehensive studies.

## Data Availability

All data produced are available online at: https://www.radc.rush.edu and ftp://ftp.sanger.ac.uk/pub/project/humgen/summary_statistics/human/2016-11-07/.

## Acknowledgements

We thank all the participants of the ROS/MAP study, the BLUEPRINT study, and the IBD GWAS and AD GWAS for their participation and generous donations.

## Author information

Center for Translational and Computational Neuroimmunology, Department of Neurology and the Taub Institute for Research on Alzheimer’s disease and the Aging brain, Columbia University Irving Medical Center, New York, NY, USA.

Lu Zeng, Charles C. White, Hans-Ulrich Klein, Philip L. De Jager

Rush Alzheimer’s Disease Center, Rush University Medical Center, Chicago, IL, USA.

David A. Bennett

## Author Contributions

LZ and PLD concept and designed the study; LZ performed the research; CCW provided data and code contributed to the analyses; HUK and DAB participated in the discussion and interpretation of the results; LZ and PLD wrote the paper; LZ created the figures which were edited by PLD; All the authors reviewed and revised the paper.

